# Review of Current Evidence of Hydroxychloroquine in Pharmacotherapy of COVID-19

**DOI:** 10.1101/2020.04.16.20068205

**Authors:** Umesh Devappa Suranagi, Harmeet Singh Rehan, Nitesh Goyal

**Affiliations:** Department of Pharmacology, Lady Hardinge Medical College & Associated Hospitals New Delhi, India

## Abstract

**Importance:** The COVID-19 Pandemic has literally left the world breathless in the chase for pharmacotherapy. With vaccine and novel drug development in early clinical trials, repurposing of existing drugs takes the center stage.

**Objective:** A potential drug discussed in global scientific community is hydroxychloroquine. We intend to systematically explore, analyze, rate the existing evidence of hydroxychloroquine in the light of published, unpublished and clinical trial data.

**Evidence review:** PubMed Ovid MEDLINE, EMBASE, Google scholar databases, pre-proof article repositories, clinical trial registries were comprehensively searched with focused question of use of hydroxychloroquine in COVID-19 patients. The literature was systematically explored as per PRISMA guidelines.

**Findings:** Total 156 articles were available as of 7^th^ May 2020; of which 11 articles of relevance were analyzed. Three *in-vitro* studies were reviewed. Two open label non-randomized trials, two open label randomized control trials, one large observational study, one follow-up study and two retrospective cohort studies were systematically analyzed and rated by oxford CEBM and GRADE framework for quality and strength of evidence. Also 27 clinical trials registered in three clinical trial registries were analyzed and summarized. Hydroxychloroquine seems to be efficient in inhibiting SARS-CoV-2 in *in-vitro* cell lines. However, there is lack of strong evidence from human studies. It was found that overall quality of available evidence ranges from ‘very low’ to ‘low’.

**Conclusions and relevance:** The *in-vitro* cell culture based data of viral inhibition does not suffice for the use of hydroxychloroquine in the patients with COVID-19. Current literature shows inadequate, low level evidence in human studies. Scarcity of safety and efficacy data warrants medical communities, health care agencies and governments across the world against the widespread use of hydroxychloroquine in COVID-19 prophylaxis and treatment, until robust evidence becomes available.

**KEY POINTS:** *Question:* What is the current evidence for use of Hydroxychloroquine in pharmacotherapy of COVID-19?

*Findings:* We electronically explored various databases and clinical trial registries and identified 11 publications and 27 clinical trials with active recruitment. The in-vitro study data demonstrates the viral inhibition by hydroxychloroquine. The clinical studies are weakly designed and conducted with insufficient reporting and significant limitations. Well designed robust clinical trials are being conducted all over the world and results of few such robust studies are expected shortly.

*Meaning:* Current evidence stands inadequate to support the use of hydroxychloroquine in pharmacotherapy of COVID-19.

## Introduction

The ongoing Coronavirus disease 2019 (COVID-19) pandemic has affected most of the countries in the world with unimagined infectious disease morbidity and mortality. As per WHO (as of 7th May, 2020), there has been a total of 3,672,238 confirmed cases and 254,045 deaths due to COVID-19 worldwide.^1^ However, no specific drug has been approved for the treatment of COVID-19 except the latest FDA emergency approval of remdesivir for use in severe cases. Recent updates indicate the vaccine quest is at least a year away. Building on experience from past Ebola and MERS pandemics, various human trials on novel pharmacotherapeutics are in progress.^2^ Drugs such as remdesivir and favipiravir are in exploratory phases of clinical trials.^3^ More than 20 other drugs such as chloroquine, hydroxychloroquine, lopinavir, ritonavir, human immunoglobulin, arbidol, oseltamivir, methylprednisolone, bevacizumab, interferons and traditional Chinese medicines are aimed at repositioning for COVID-19 treatment.^4^ Forerunners among these are antimalarial drugs chloroquine and hydroxychloroquine, used extensively in treatment of malaria and elsewhere since many decades.^5,6^ These drugs are 4-aminoquinoline derivatives exhibiting wide range of in-vitro activity against viruses. Their antiviral efficacy has been attributed to many different mechanisms.^7^ Chloroquine is known to possess considerable broad-spectrum antiviral effects by interfering with the fusion process of viruses by increasing the local pH.^8^Other mechanisms include raise in endosomal pH in host cells thereby inhibiting auto-lysosome fusion and disrupting the enzymes needed for the viral replication.^9,10^

Hydroxychloroquine (HCQ) is synthesized by N-hydroxyethyl side chain substitution of chloroquine. Although the antimalarial activity of HCQ is equivalent to that of chloroquine, HCQ is preferred over chloroquine owing to its lower ocular toxicity.^11^It is also used in the treatment of rheumatoid arthritis, chronic discoid lupus erythematosus, and systemic lupus erythematosus. In addition to endosomal pH increase, HCQ is also said to inhibit terminal glycosylation of ACE2 receptor, considered as target of SARS-CoV and SARS-CoV-2 cell entry.^12^The non-glycosylated ACE2 receptor might interact inefficiently with the SARS-CoV-2 spike protein, thus inhibiting the viral entry.^13^These myriad mechanisms of HCQ and its relative lesser toxicity profile as compared to chloroquine make it an attractive candidate in the pursuit of drug repositioning. In this highly demanding scenario of unmet need and steeply increasing morbidity and mortality of COVID-19, many government bodies and expert panels have recommended the use of chloroquine and HCQ for prophylaxis and treatment of COVID-19.^14-18^ In such situation of urgency, there is a need to explore the current literature and critically analyze the existing evidence. We intend to conduct a detailed systematic search analysis of current literature and propose our findings.

## Materials and methods

### Data sources

A comprehensive literature search was done independently by each author to find the role of HCQ in COVID-19 disease. PubMed Ovid MEDLINE, EMBASE, Google scholar databases were searched for existing literature from 2019 to 7^th^ May 2020, 2020. The clinical trial Registries of the United States (clinicaltrials.gov), Chinese Clinical Trial Registry, WHO International clinical trial registry platform (ICTRP) were searched for ongoing registered studies. For preprint/pre-proof articles, repositories like BioRxiv, MedRxiv and ChemRxiv were searched.

### Literature search

Search words included MeSH Terms (hydroxychloroquine OR HCQ) AND (COVID-19 OR Coronavirus OR nCov2 OR SARS-CoV2). We searched for both published and unpublished studies extensively. No language, time, study type and demographic filters were used. The search expansion was done using a snowballing method applied to the authors and references of selected publications. PRISMA guidelines were followed. Article search included abstracts, original research, *in-vitro* experimental studies, observational studies and controlled/uncontrolled trials. We excluded the articles like news items, magazine pieces, duplicate papers, review articles, editorials and letters to editor, expert opinions, perspectives, consensus statements and articles without the mention of the role of HCQ in COVID-19 or HCQ use in other conditions.

We searched databases of clinical trial registries using the search terms ‘Hydroxychloroquine’, ‘HCQ’, ‘Plaquenil’, ‘COVID-19’, ‘SARS-CoV2’, ‘novel Corona virus’ ‘nCoV 2’. After identification and elimination of duplicated appearances, 96 clinical trials were found to be registered. Each database was further scanned and analyzed to remove the non-recruiting, inactive and cancelled trials, finally yielding 27 randomized control trials (RCTs) currently undergoing active recruitment for COVID-19 treatment with HCQ.

### Screening, data extraction, data analysis, critical appraisal and evidence rating

Screening of articles was done independently by investigators according to titles, abstracts, summaries and conclusions. Methodical data extraction was done from selected articles and pertinent portions were identified, tabulated and presented systematically in the form of tables &summary. Randomized clinical trials with active recruitment were analyzed after collecting publically available information on various clinical trial databases. We used GRADE (Grading of Recommendations, Assessment, Development and Evaluations) framework methodology to rate the certainty of evidence from both published and unpublished clinical studies and Oxford center for evidence based medicine (CEBM) to assess and rate the quality of evidence.

## Results

Total 156 articles were identified on initial search of databases. Following screening of titles and abstracts and removal of duplicates, eleven articles (three *in-vitro* studies, two open label non-randomized trials, two open label randomized control trials, one large observational study, one follow-up study, two retrospective cohort studies) were selected for further data extraction and analyses. Out of these 11, four clinical studies (one open label randomized control trial, one open label non-randomized trial and two retrospective cohort studies) were from pre-print servers. We also identified 96 clinical trials registered in three clinical trial registry databases. Methodical screening and analysis further yielded 27 RCTs currently undergoing active recruitment.

**Figure1:**
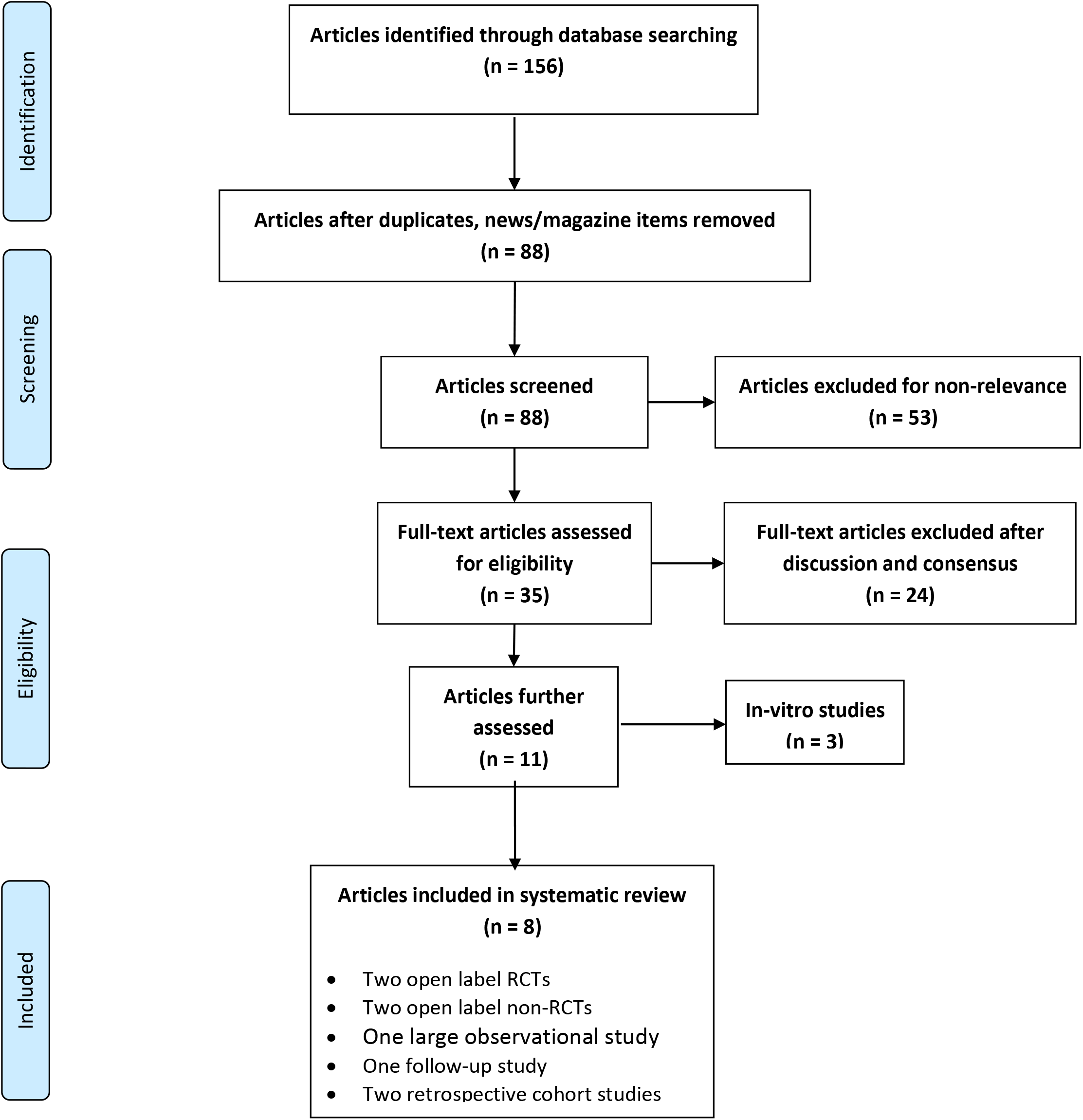
PRISMA flow diagram.

### *In vitro* studies of Hydroxychloroquine demonstrating anti-coronaviral activity

Yao et al, assessed the pharmacological activity of chloroquine and HCQ using SARS-CoV-2 infected Vero cells. Further as continued part of the study, they simulated physiologically-based pharmacokinetic models (PBPK) on the *in vitro* data obtained. The researchers found HCQ to be more potent than chloroquine to inhibit SARS -CoV-2 *in vitro.* Based on PBPK extrapolation, they recommended a loading dose of 400 mg twice daily of HCQ sulfate given orally, followed by a maintenance dose of 200 mg given twice daily for 4 days.^19^

In another correspondence report letter of an *in vitro* study by Liu et al, the investigators used VeroE6 cells and compared the antiviral activity of chloroquine versus HCQ against SARS-CoV-2 to determine different multiplicities of infection (MOIs) by quantification of viral RNA copy numbers. They found out that 50% maximal effective concentration (EC50) for HCQ was significantly higher than chloroquine and HCQ can efficiently inhibit SARS-CoV-2 infection *in vitro.^20^*

Previously in 2006, French researchers demonstrated that chloroquine and HCQ effectively inhibit both human and feline SARS COV in the infected Vero cells. EC50 for HCQ was significantly higher than chloroquine.^21^ (Table 1)

**Table 1:**
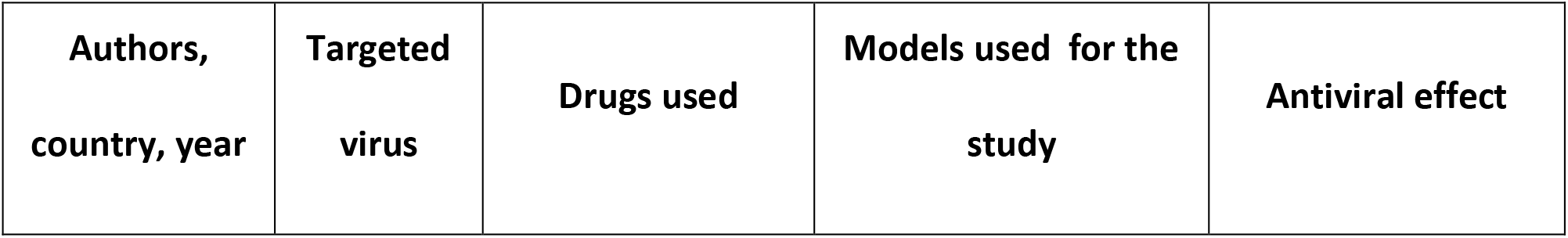
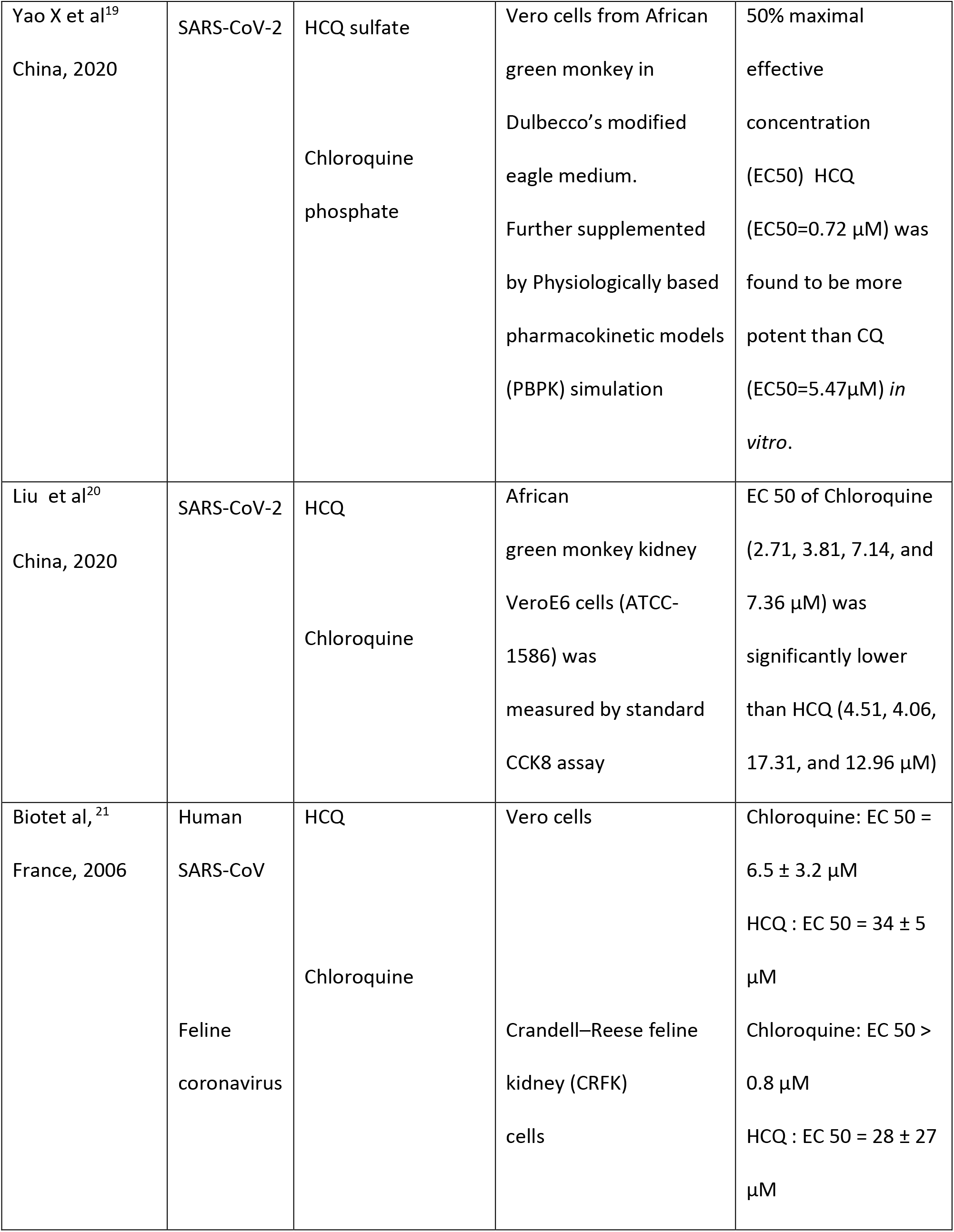
Summary of *in-vitro* studies showing efficacy of hydroxychloroquine against SARS-CoV-2 infected Cell lines.

| **Authors,**<br>**country, year** | **Targeted**<br>**virus** | **Drugs used** | **Models used for the**<br>**study** | **Antiviral effect** |
| --- | --- | --- | --- | --- |
| Yao X et al<sup>19</sup><br>China, 2020 | SARS-CoV-2 | HCQ sulfate<br>Chloroquine<br>phosphate | Vero cells from African<br>green monkey in<br>Dulbecco's modified<br>eagle medium.<br>Further supplemented<br>by Physiologically based<br>pharmacokinetic models<br>(PBPK) simulation | 50% maximal<br>effective<br>concentration<br>(EC<sub>50</sub>) HCQ<br>(EC<sub>50</sub>=0.72 µM) was<br>found to be more<br>potent than CQ<br>(EC<sub>50</sub>=5.47µM) *in*<br>*vitro*. |
| Liu et al<sup>20</sup><br>China, 2020 | SARS-CoV-2 | HCQ<br>Chloroquine | African<br>green monkey kidney<br>VeroE6 cells (ATCC-1586) was<br>measured by standard<br>CCK8 assay | EC 50 of Chloroquine<br>(2.71, 3.81, 7.14, and<br>7.36 µM) was<br>significantly lower<br>than HCQ (4.51, 4.06,<br>17.31, and 12.96 µM) |
| Biotet al,<sup>21</sup><br>France, 2006 | Human<br>SARS-CoV<br>Feline<br>coronavirus | HCQ<br>Chloroquine | Vero cells<br>Crandell–Reese feline<br>kidney (CRFK)<br>cells | Chloroquine: EC 50 =<br>6.5 ± 3.2 µM<br>HCQ : EC 50 = 34 ± 5<br>µM<br>Chloroquine: EC 50 ><br>0.8 µM<br>HCQ : EC 50 = 28 ± 27<br>µM |

### Clinical studies conducted in COVID-19 patients

In the positive background of successful *in vitro* data and in the situation of an emerging epidemic, the Chinese authorities issued a consensus statement for the use of chloroquine in COVID-19 patients.^14^The earliest data of chloroquine administration in humans came from various parts of China in the form of collective reports were published by Gao et al.^22^ The authors reported clinical experience data of treating more than 100 patients with chloroquine in various locations. They mentioned that chloroquine reduced the duration of illness and improved the pneumonia and pulmonary image changes in COVID-19 positive patients. The authors also recommended the drug to be included in the COVID-19 Guidelines issued by the National Health Commission of China for the use of drug in larger populations.^23^

The first empirical evidence of use of HCQ in humans was obtained by a small RCT conducted by Chen et al^24^ in 30 adult COVID-19 patients. The treatment group received 400mg HCQ for 5 days, while the standard care was given to control group. The primary outcome was nasopharyngeal swab test results on Day 7. Investigators found that there is no difference between treatment and control group in the number of patients testing negative for COVID-19 on Day 7 (13 v/s 14), the duration of illness did not differ significantly (p= >0.05). There was one drop out and seven (three in treatment group and four in control) adverse events. The authors concluded that COVID-19 has good prognosis and larger sample size with better endpoints is needed to investigate the effects further.^24^

An open-label, non-randomized clinical trial was conducted by Gautret et al^25^ in France with 36 patients diagnosed with COVID-19. HCQ in dose of 200mg three times daily was given to 20 patients for 10 days, additionally six patients in this group received azithromycin (500 mg on day 1, 250mg on days 2-5) to prevent bacterial superinfection. The control group received standard care. The primary outcome was detection of SARS-CoV-2 RNA in nasopharyngeal samples. The authors reported that patients in the treatment group significantly differed for SARS-CoV-2 detection than controls. On Day 6 of post initiation, 70% of HCQ treated patients were virologically cured compared to 12.5% in the control group (p= 0.001). They concluded that HCQ treatment is significantly associated with viral load reduction/disappearance in COVID-19 patients and its effect is reinforced by azithromycin.^25^

A six-day pilot, uncontrolled, non-comparative observational follow-up study was conducted by French investigators to assess the clinical and microbiological effect of a combination of HCQ and azithromycin in 80 COVID-19 patients. The investigators reported that all patients but two (a 86 years old succumbed to illness, a 74 years old needed ICU) showed clinical improvement with the combination therapy. qPCR testing showed a rapid fall of nasopharyngeal viral load- 83% and 93% patients were negative at Day 7, and Day 8 respectively. 97.5% of respiratory samples were negative for virus cultures at Day 5. The researchers urged to evaluate the combination strategy to treat patients in early course and avoid the spread of the disease.^26^

A latest large observational study conducted by American researchers, assessed the association between HCQ use and intubation or death. 1376 patients were followed up for median of 22.5 days. Among them, 811 (58.9%) patients received HCQ (600 mg twice on day 1, then 400 mg daily for a median of 5 days). Overall 346 patients (25.1%) had a primary end-point event (180 patients were intubated, of whom 66 subsequently succumbed, and 166 died without intubation). The authors reported that there was no significant association between HCQ use and intubation or death. They concluded hydroxychloroquine administration was not associated with either a greatly lowered or an increased risk of the composite end point of intubation or death.^27^ (Table 2)

**Table 2:**
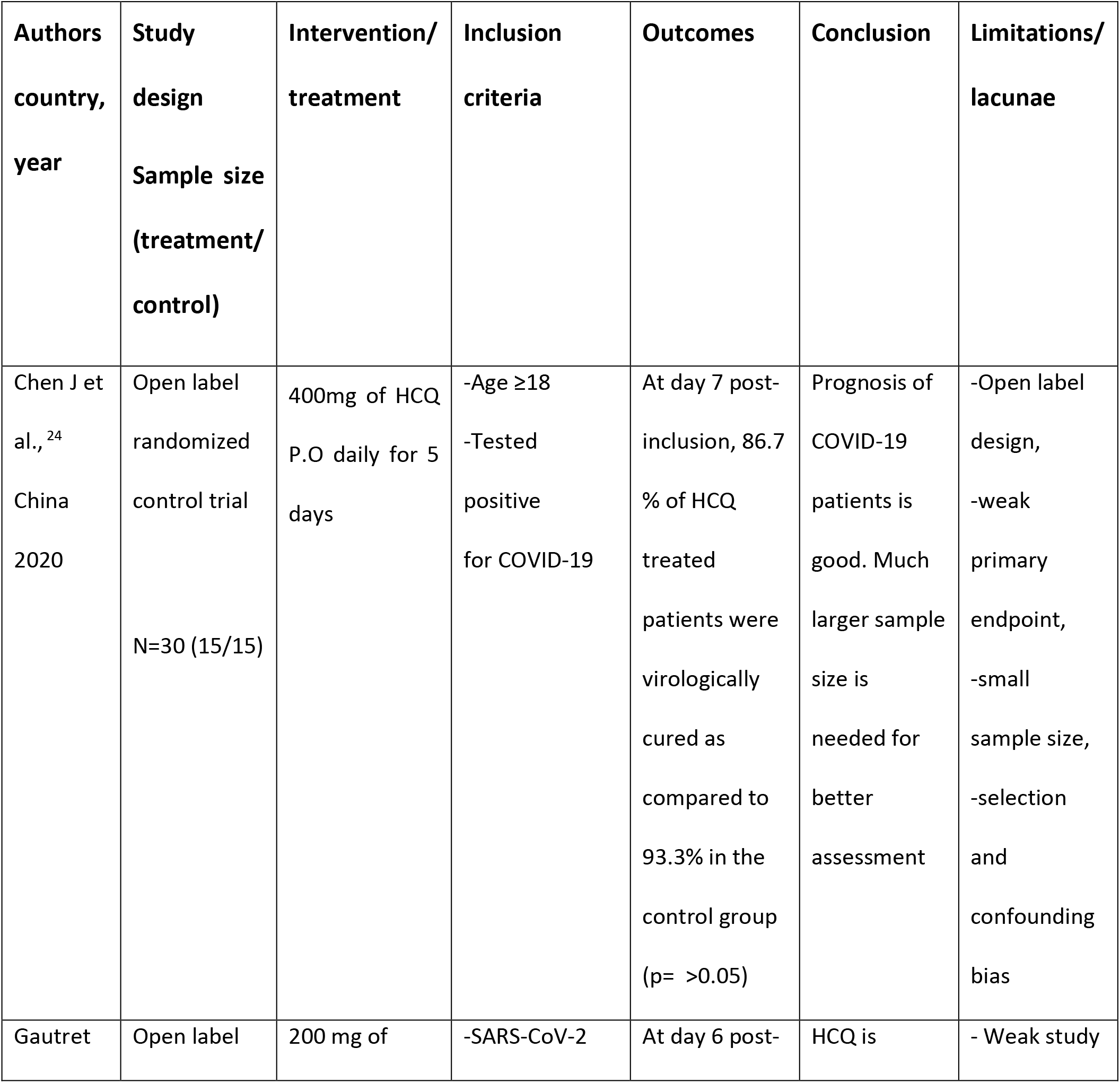
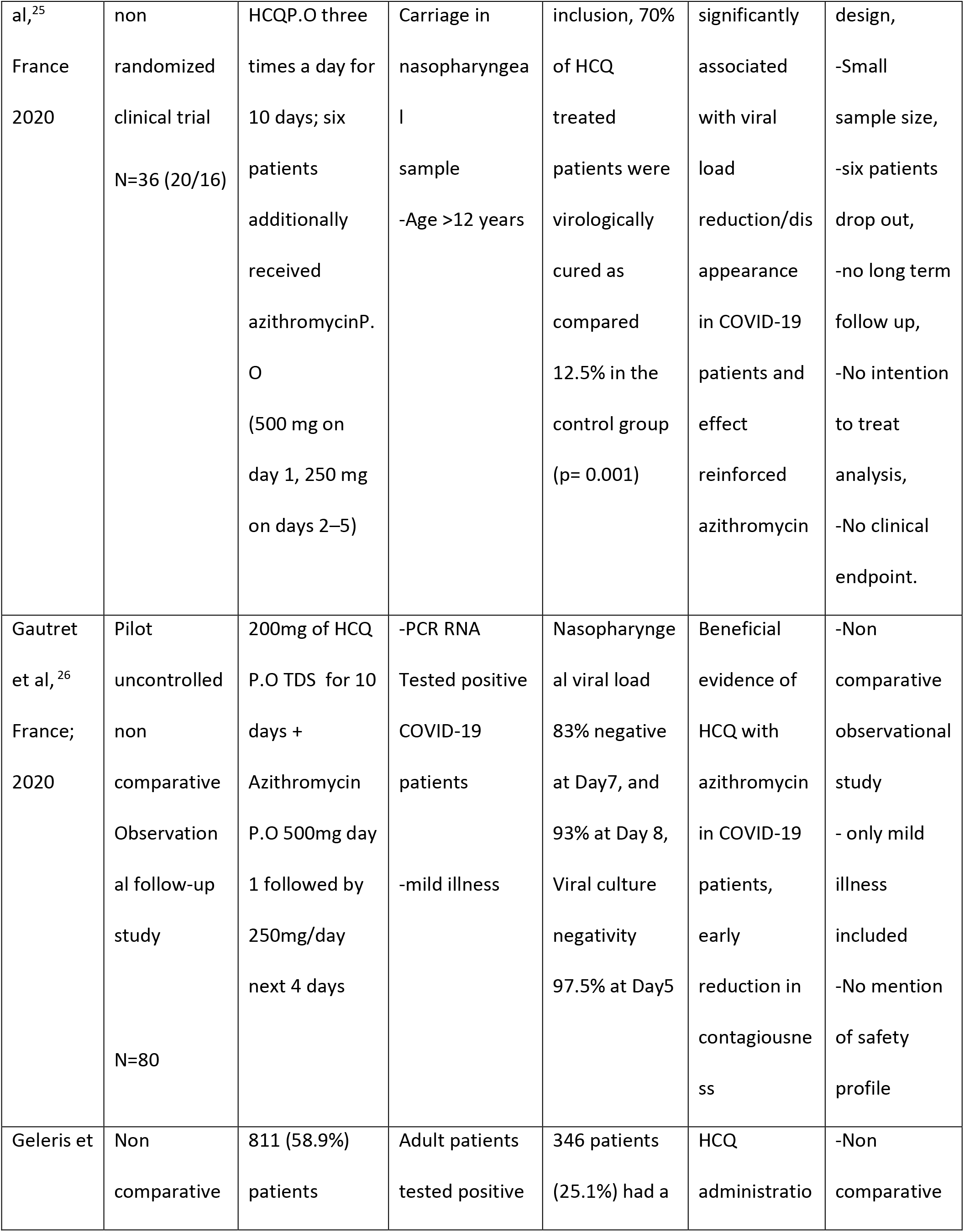
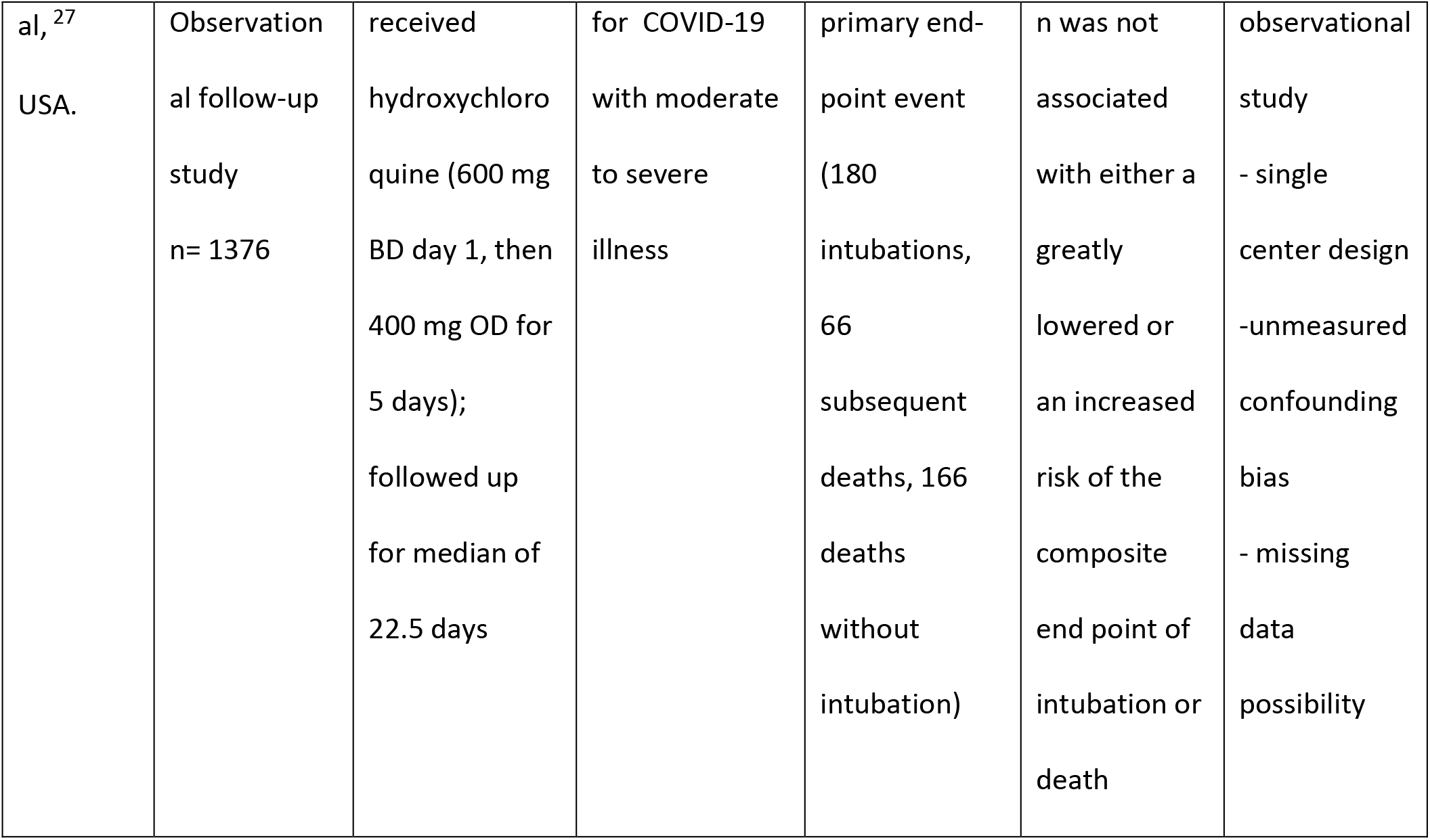
Summary of clinical studies with hydroxychloroquine treatment in COVID-19 patients.

### Unpublished studies from preprint repositories

We searched preprint servers for pre-proof, unpublished, approval awaited studies and articles. Since these studies are yet to be peer-reviewed we have briefly summarized their findings along with the limitations and lacunae. (Table 3)

**Table 3:**
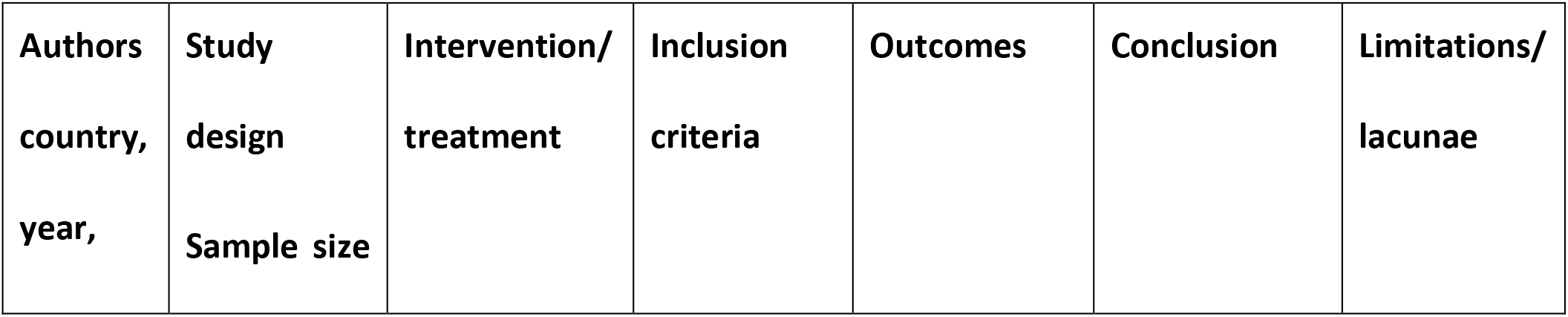
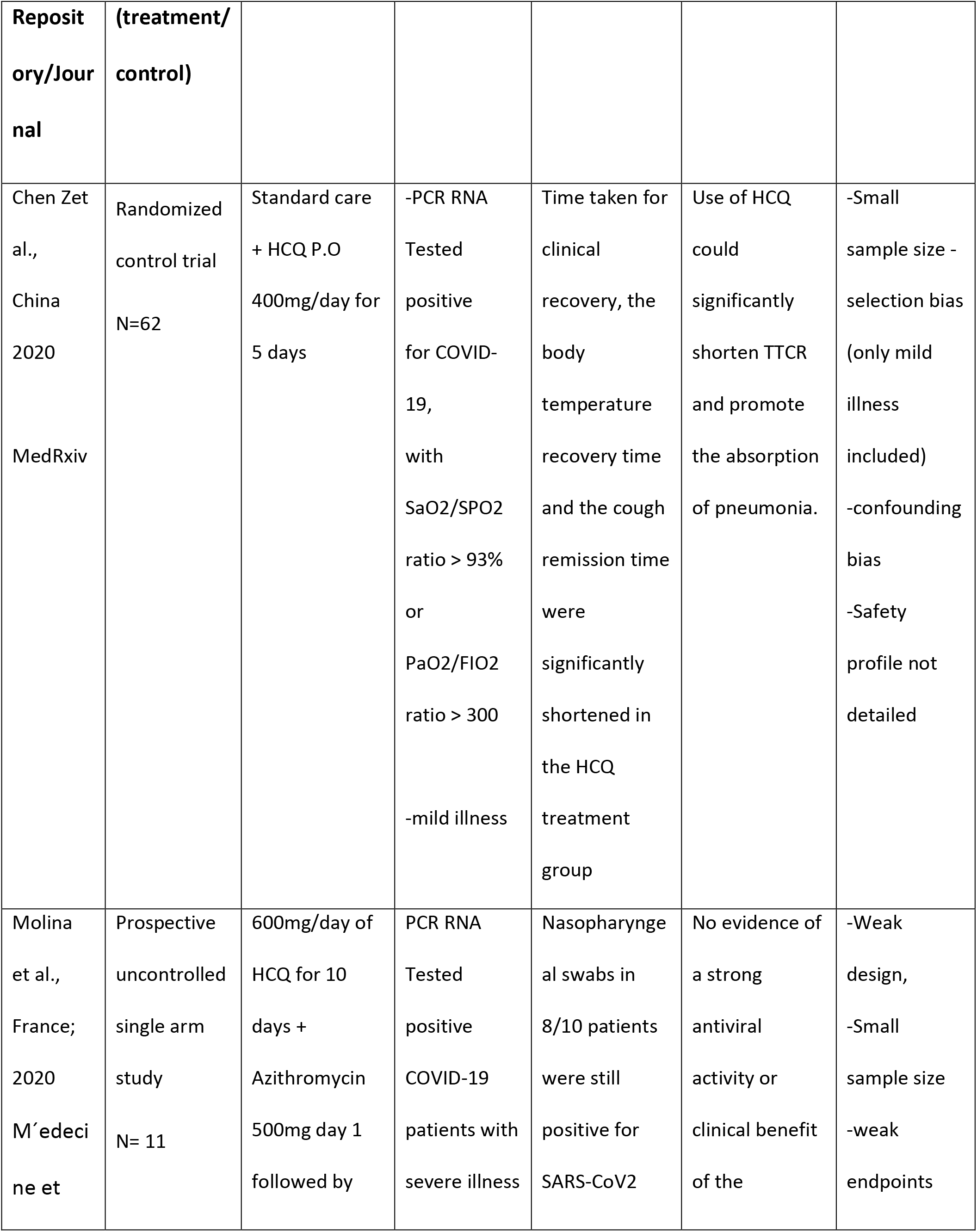
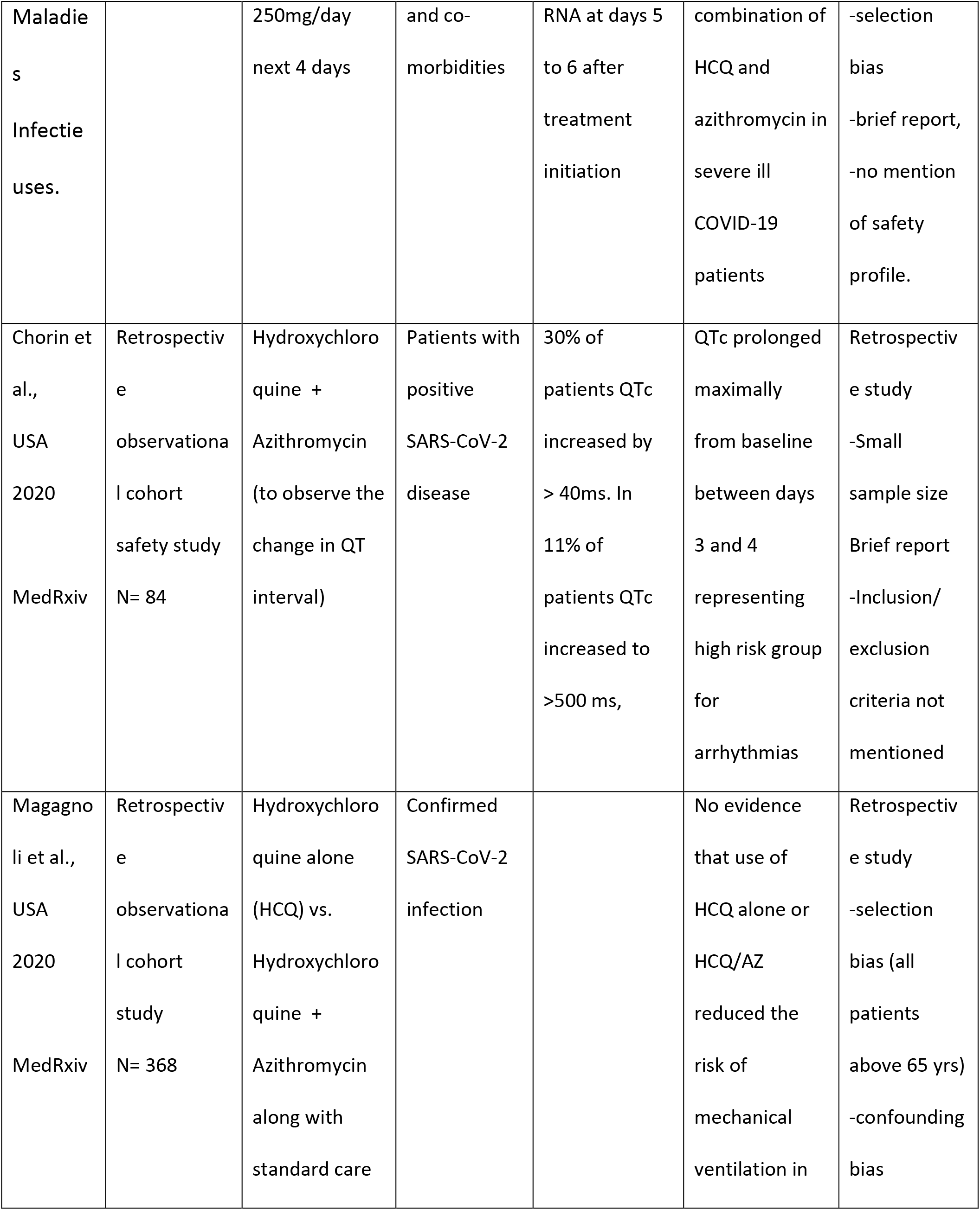
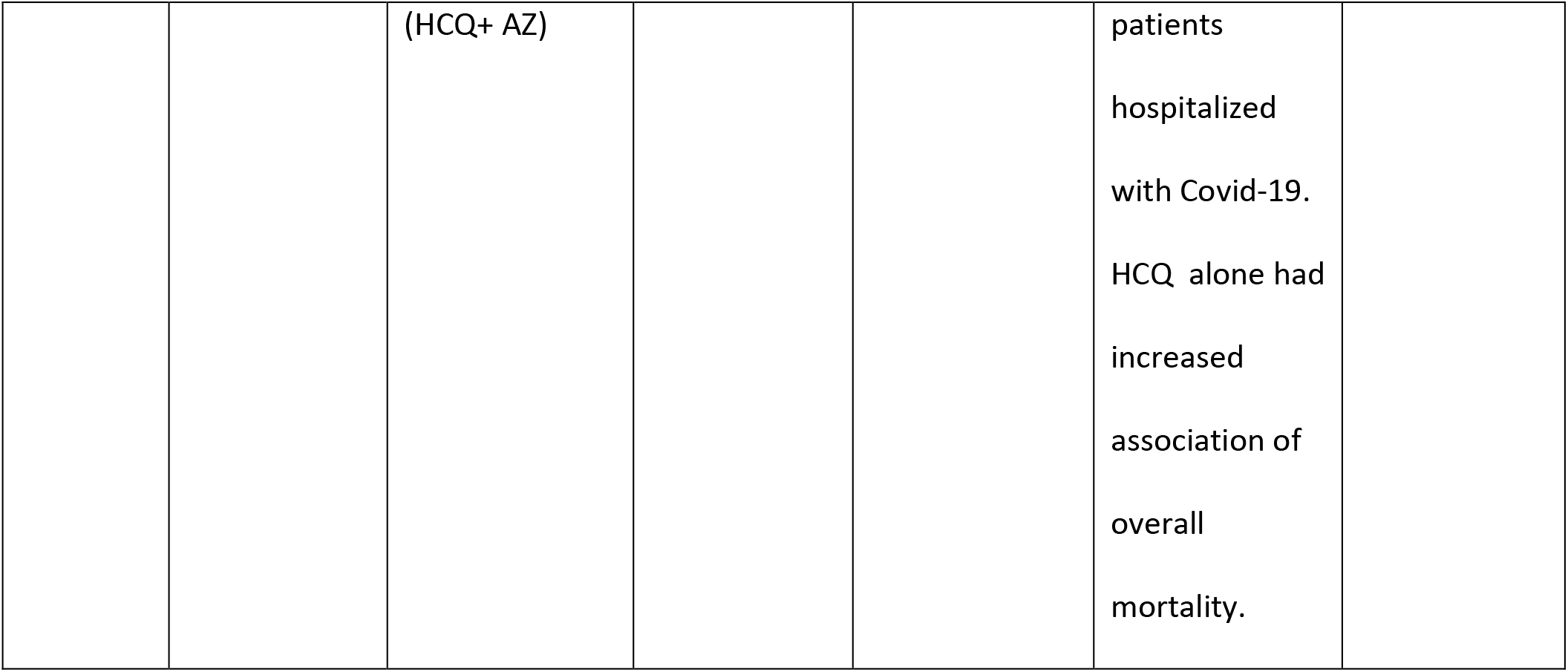
Summary of unpublished studies reporting the use of hydroxychloroquine in treatment of COVID-19 patients.

### Assessment of methodological quality and rating of evidence generated by clinical studies

We used GRADE framework approach (Figure 2) to assess the methodological quality of published and unpublished clinical studies of HCQ in COVID-19. The Oxford center for evidence based medicine (CEBM) levels of evidence was used to assess and rate the quality of evidence. Individual outcomes, overall outcome and clinical relevance were applied to rate the strength and quality of evidence. (Table 4)

**Figure 2:**
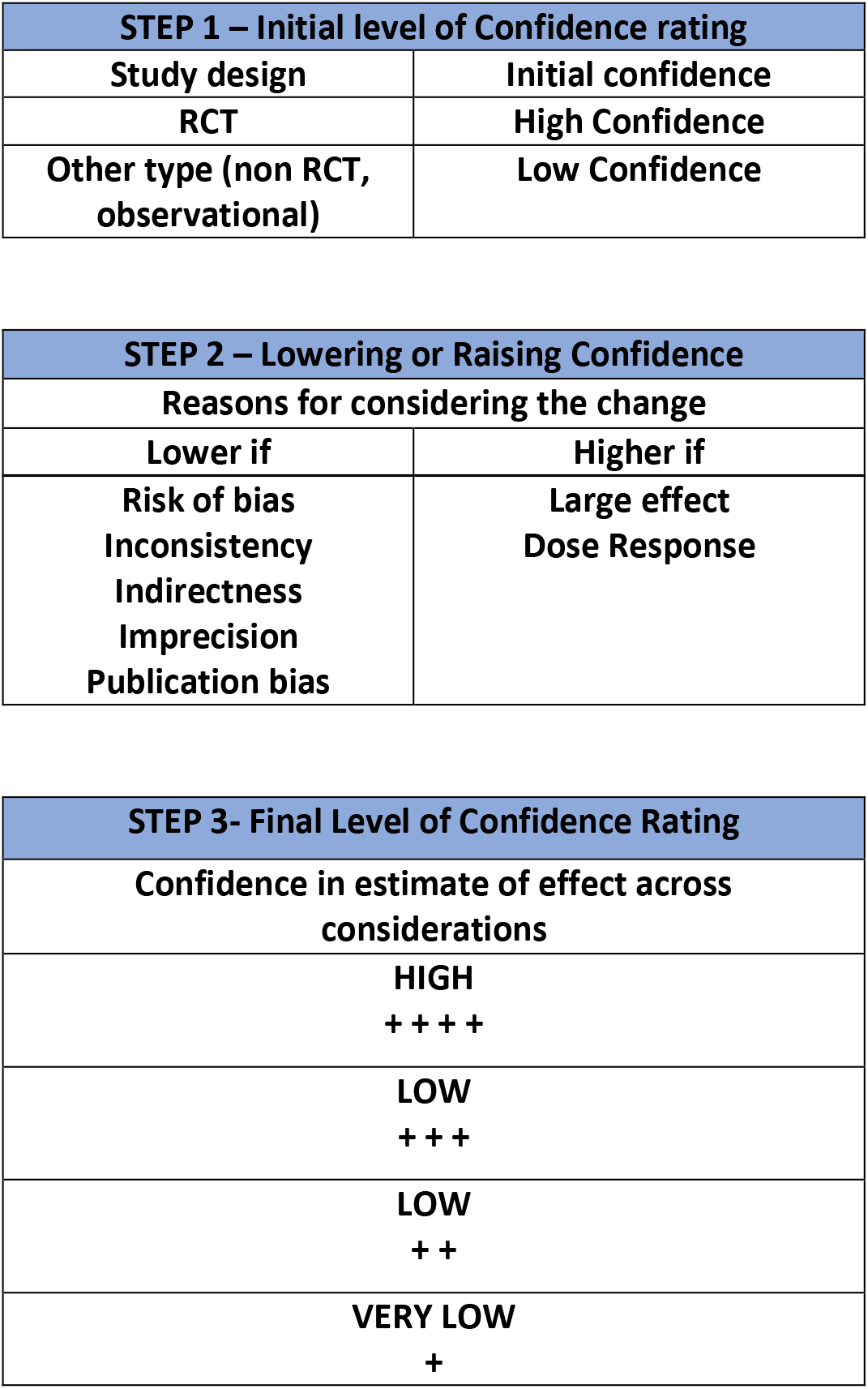
Approach to rating of quality of evidence using GRADE methodology.

**Table 4:**
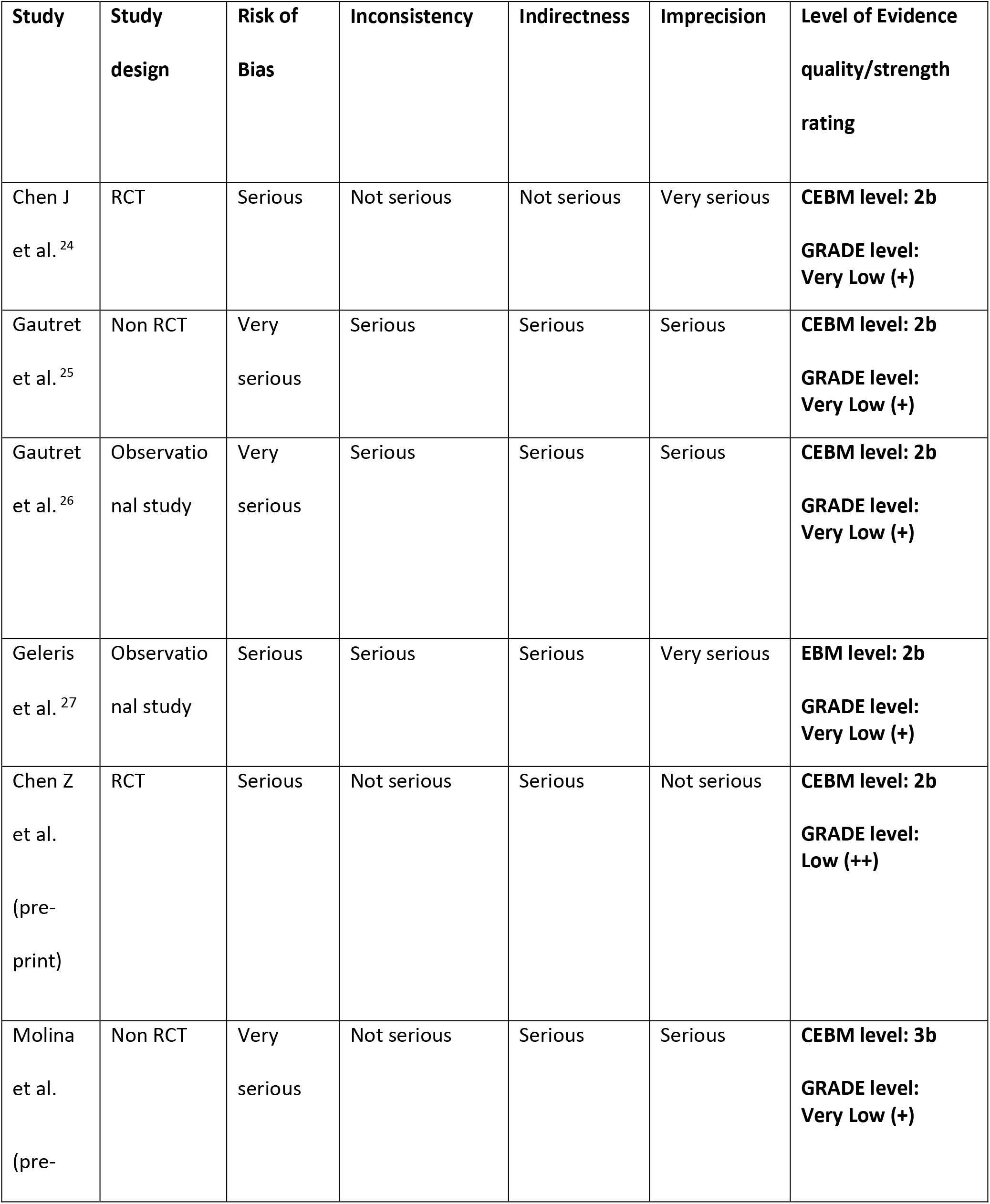
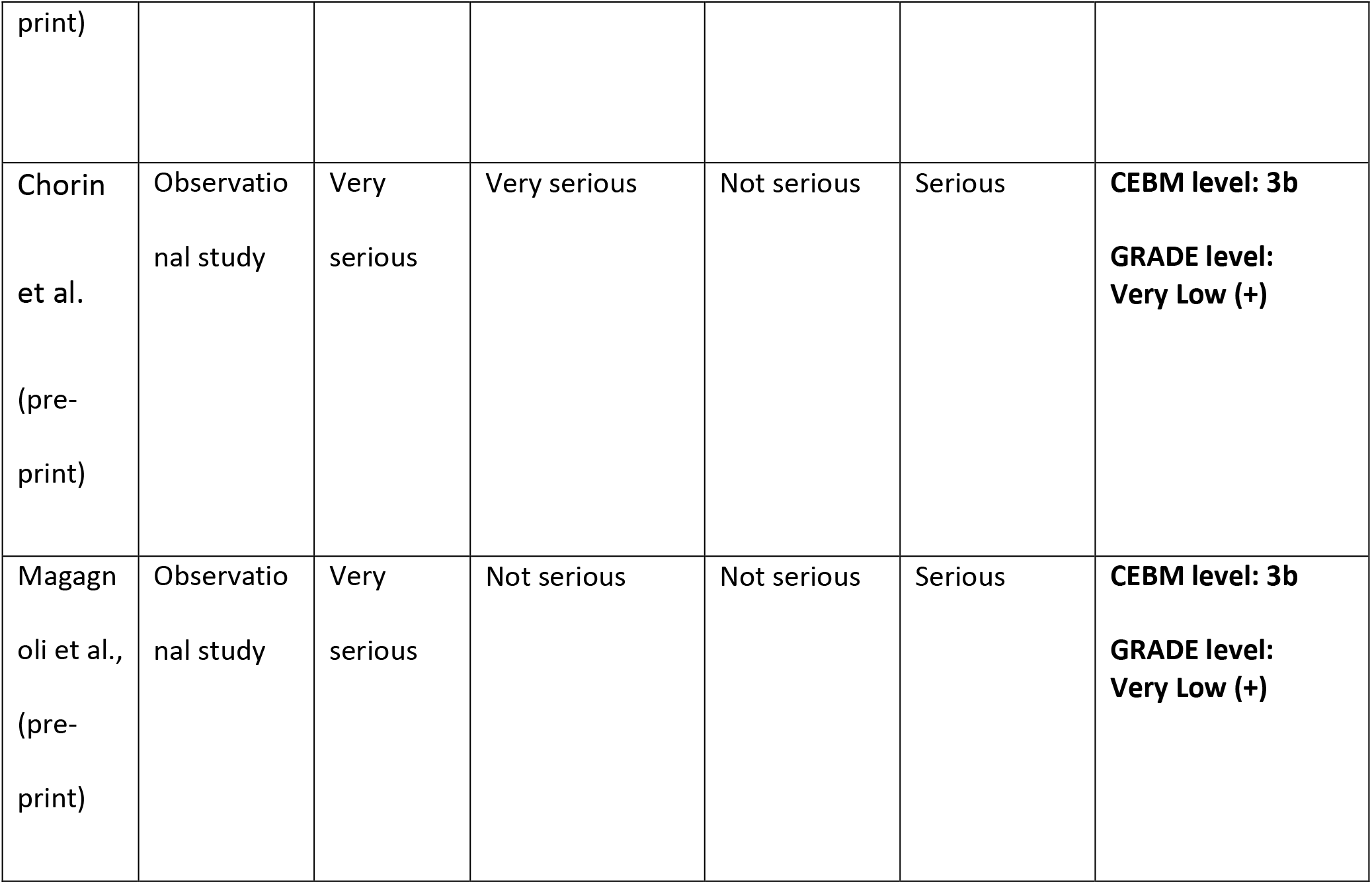
Summary of assessment of quality and strength of evidence of clinical studies.

### Summary of Ongoing Clinical trial data from clinical trial registry databases

Many of the ongoing RCTs conducted are studying the effect of HCQ compared to placebo(NCT04342221, NCT04333654, NCT04332991, NCT04331834), few of the RCTs have parallel design arms of HCQ and azithromycin (NCT04341727,NCT04341207, NCT04334382). Some RCTs have robust trial designs with quadruple masking and strong endpoints (NCT04333654, NCT04332991,NCT04331834). Few RCTs are in advance phases of clinical trials (NCT04316377, NCT04341493) and some studies have large sample size to measure the effect with higher strength of confidence (NCT04328012, NCT04328467). There are studies which are considering the safety endpoints in the main outcome measures (ChiCTR2000029868). Some RCTs are also testing antiretroviral drugs like lopinavir/ritonavir, emtricitabine/tenofovir along with HCQ arm (NCT04328012,NCT04334928). Other studies are interested in tocilizumab (NCT04332094) and umefenovir/arbidol (ChiCTR2000029803) along with HCQ. Few of the studies are being conducted in severely ill patients (NCT04325893, ChiCTR2000029898) and some in mild infections of COVID-19 (NCT04307693, ChiCTR2000029899). Most of the studies are registered to be conducted in United States and China, others being conducted in Spain (NCT04331834, NCT04332094), Norway (NCT04316377), France (NCT04325893, NCT04341207), Germany (NCT04342221), Denmark (NCT04322396), Brazil (NCT04322123, NCT04321278), Mexico (NCT04341493) and Republic of Korea (NCT04307693). Few earlier studies registered in China (in Feb 2020) are nearing their completion in April end or early May 2020, their results can be expected in near future (ChiCTR2000029898, ChiCTR2000029899, ChiCTR2000029992). Additional details regarding the ongoing trials can be obtained from Supplementary Table 1.

In Europe, the Discovery project (NCT04315948) study has commenced in late march 2020 and with recruitment of 3100 patients. The four treatments set to be evaluated in the discovery project as per WHO recommendations are Remdesivir, Lopinavir/Ritonavir, IFN0-1a, Hydroxychloroquine/Chloroquine. The first set of results is expected to be available in 3 to 4 weeks of time. The estimated study completion date has been set in March 2023.^28^

## Discussion

As on 7^th^ May 2020, COVID-19 pandemic has caused more than three and half million infections and more than 250,000 deaths growing up in alarming rate. Specific pharmacotherapy is the highest need of the world. Hydroxycholoroquine with relatively better safety profile than choloroquine and possible better antiviral efficacy^19^ offers a compelling hope. We systematically searched various databases and clinical trial registries to evaluate the evidence. During the previous outbreak of SARS, an *in vitro* study demonstrated the anti-corona viral effect of HCQ and choloroquine.^21^ More recently Chinese researchers conducted in-vitro studies in cell lines and demonstrated the potential antiviral activity of HCQ against SARS-CoV2 as compared to chloroquine.^19,20^ It is relevant to note that these studies were the basis of initial opinions and general consensus statements given by various panels across the world during the early stages of the pandemic. We found out that there is scarcity of well conducted and adequately reported human studies of HCQ use in COVID-19. This is in agreement with the other authors with similar findings of lack of literature in this regard.^29-32^ Literature also lacks studies conducted in healthcare workers for either prophylaxis or treatment. Gao et al reported more than 100 patients with COVID-19 pneumonia showed clinical improvement and changes in image findings on chloroquine administration.^22^ It is pertinent to note that this letter was the brief report of ongoing many trials in various locations in China, neither it mentioned any specific data regarding interventions, study design, study population and outcome measures, nor any adverse events were discussed. Chen et al in a RCT involving 30 COVID-19 patients did not find any significant difference between treatment and control group in both nasopharyngeal swab negativity and duration of illness.^24^ This study was an open label trial with small sample size and had high risk of confounding and selection bias, the authors agreed that primary end point was weak and more robust end points with larger sample size is required to establish the effects.

Gautret et al^25^ in a non-randomized clinical trial in 36 COVID-19 patients, reported viral load reduction by HCQ and its reinforcement by azithromycin. This study had major limitations in the form of small sample size, absence of randomization and masking, lack of intention to treat analysis and long term follow up, there was no clinical endpoint as outcome measure. A follow up study by Gautret et al in 80 COVID-19 patients reported 97.5% of respiratory samples were negative for virus cultures at Day 5. This study too involved only mild illness patients and did not report adverse effect profile and being an uncontrolled observational study, the strength of evidence tends to be low.^26^ A latest large American observational study in 1376 patients reported no association between hydroxychloroquine administration and intubation/death in moderate to severe COVID-19 patients. This study had limitations of single center observational design, confounding bias and possibility of missing data events.^27^

We assessed the methodological quality and certainty of evidence of both published and unpublished clinical studies in existing literature and found that overall quality of available evidence ranges from ‘very low’ to ‘low’; the Oxford CEBM rating used showed the quality of studies to be mostly at 3b level and couple studies at 2b level. (Table 4)

We also searched, identified and analyzed clinical trial databases to explore the ongoing active clinical trials (Supplementary Table 1) and found out relevant 27 clinical trials. Few trials among these are in advanced phases. Earlier registered Chinese clinical trials are expected to report the results in near future and robust designed RCTs elsewhere in the world are expected to produce their interim findings shortly henceforth.

It is appropriate to note that none of the available studies of HCQ in COVID-19 have emphasized on the adverse effects and toxicity profile of the drugs in treated patients. Even though HCQ has relatively better safety profile than chloroquine, owing to its prolonged pharmacokinetics (537 hours half-life) and gradual elimination, HCQ has potential to cause various adverse events viz. gastrointestinal upset,^33^ retinal toxicity,^34^ fulminant hepatic failure,^35^ severe cutaneous adverse reactions.^36^ An important adverse effect of HCQ is cardiac conduction defects and ventricular arrhythmias. QT prolongation and arrhythmias can be precipitated by concomitant use of azithromycin.^37^ Small but absolute risk of cardiovascular death is seen to be associated significantly with azithromycin as compared to fluoroquinolones.^38^ Overdose or poisoning of HCQ is difficult to treat, caution is warranted in patients with hepatic and renal dysfunction, and regular ECG monitoring is advised in patients with cardiovascular diseases and in electrolyte imbalances.^39^ Irrational use in general population without credible evidence may pose greater risk than benefit.

To best of our knowledge, this systematic review is the most comprehensive exploration and analysis of existing literature in this topic till date. Our systematic review has limitations in its rigor due to the inadequate, inconsistent data and heterogeneity of studies available. The rapidly expanding knowledge base of COVID-19 poses the possibility that some studies remain un-captured. However, we have tried our best to mitigate this by allowing broad, flexible search terms and by including many databases and preprint repositories, while remaining focused on the research question. In this background, we believe that expert opinions and clinical consensus statements given by various international authorities for the use of HCQ either as prophylaxis to high risk individuals ^15^ and healthcare professionals^16^ or as emergency treatment of COVID-19 patients^17,18^ lack strong evidence base.

## Conclusion

The in-vitro cell culture based data of viral inhibition does not suffice for the use of hydroxychloroquine in the patients with COVID-19. Current literature shows scant and low level evidence in clinical studies. At this stage it is reasonable to suggest against the use hydroxychloroquine as prophylaxis both in general population as well as health care workers. Considering the toxicity profile, chances of overdoses and poisoning can pose serious health threats if hydroxychloroquine is used widely. Ongoing well designed clinical trials are expected to provide explicit answer on safety and efficacy in near future. It is warranted against the widespread use of hydroxychloroquine in COVID-19 until robust evidence becomes available.

## Data Availability

All data referred to in the manuscript are available with the authors. we assert and agree for the same.

---

**Supplementary table 1.** Ongoing Clinical trial data from clinical trial registry databases.

Ongoing Clinical trial data from clinical trial registry databases
| Sl. no | Title | Trial Reg number | Intervention | Comparator | Study Design | Main outcome(s) | Population | Place/ Country | Expected Timeline |
| --- | --- | --- | --- | --- | --- | --- | --- | --- | --- |
| 01 | Hydroxychloroquine for COVID-19 Study | NCT04342221 | Hydroxychloroquine Sulfate | Placebo | Phase 3 RCT, quadruple masking | Effect of HCQ on in vivo viral clearance | N= 220<br>18 Yrs to 99 Yrs (Adult, Older Adult) All sex | Germany | Start- Mar 29, 2020<br>End- Mar 2021 |
| 02 | Hydroxychloroquine, Azithromycin in the Treatment of SARS CoV-2 Infection (WU352) | NCT04341727 | Hydroxychloroquine Sulfate<br>Azithromycin<br>Chloroquine sulfate | - | Open label RCT, parallel design | Hours to recovery, Time fever resolution | N= 500<br>18 Yrs (Adult, Older Adult) All sex | USA | Start- Apr 4, 2020<br>End- Apr 1 2021 |
| 03 | Hydroxychloroquine vs Nitazoxanide in Patients With COVID-19 Study | NCT04341493 | Nitazoxanide 500 mg<br>Hydroxychloroquine | - | Phase 4 RCT, single masking, parallel design | Mechanical ventilation requirement | N= 86<br>5 Yrs and older (child Adult, Older Adult) All sex | Mexico | Start- Apr 6, 2020<br>End- Aug 30, 2020 |
| 04 | Epidemiology of SARS-CoV-2 and Mortality to Covid19 Disease in French Cancer Patients Study | NCT04341207 | Hydroxychloroquine<br>Azithromycin | - | Phase 2 , non randomised Open label trial | Prevalence and the 3-months incidence of SARS-CoV-2 in cancer patients | N= 1000<br>18 Yrs and older(Adult, Older Adult) Both sex | France | Start- Apr 3, 2020<br>End- Apr2022 |
| 05 | Hydroxychloroquine vs. Azithromycin for Outpatients in Utah With COVID-19 Study | NCT04334382 | Hydroxychloroquine<br>Azithromycin | - | Phase 3 Open label, parallel design RCT | Hospitalization within 14 days of enrolment, Duration of COVID-19-attributable symptoms | N= 1550<br>45 Yrs and older(Adult, Older Adult) All sex | USA | Start- Apr 2, 2020<br>End- Dec 31 2021 |
| 06 | Hydroxychloroquine in Outpatient Adults With COVID-19 Study | NCT04333654 | Hydroxychloroquine SAR321068 | Placebo | Phase 1 RCT, parallel design, quadruple masking | Change from baseline to Day 3 in nasopharyngeal SARS-CoV-2 viral load | N= 210<br>18 Yrs and older(Adult, Older Adult) All sex | USA | Start- Mar 31, 2020<br>End- May 2020 |
| 07 | Outcomes Related to COVID-19 Treated With Hydroxychloroquine Among In-patients With Symptomatic Disease Study | NCT04332991 | Hydroxychloroquine | Placebo | Phase 3 RCT, parallel design, quadruple masking | COVID Ordinal Outcomes Scale on Day 15, all-location, all-cause mortality assessed on day 15 | N= 510<br>18 Yrs and older(Adult, Older Adult)<br>All sex | USA | Start- Apr, 2020<br>End- Apr 2021 |
| 08 | Clinical Trial of Combined Use of Hydroxychloroquine, Azithromycin, and Tocilizumab for the Treatment of COVID-19 Study | NCT04332094 | Tocilizumab<br>Hydroxychloroquine<br>Azithromycin | - | Phase 2 Open label, parallel design RCT | In-hospital mortality<br>Need for mechanical ventilation in ICU | N= 276<br>18 Yrs and older (Adult, Older Adult)<br>All sex | Spain | Start- Apr, 2020<br>End- Sep 2020 |
| 09 | Pre-Exposure Prophylaxis With Hydroxychloroquine for HighRisk Healthcare Workers During the COVID-19 Pandemic Study | NCT04331834 | Hydroxychloroquine | Placebo | Phase 3 RCT, parallel design, quadruple masking | Confirmed cases of a COVID-19, SARS-CoV-2 seroconversion | N= 440<br>18 Yrs and older (Adult, Older Adult)<br>All sex | Spain | Start- Apr 3, 2020<br>End-Oct 3, 2020 |
| 10 | Hydroxychloroquine vs. Azithromycin for Hospitalized Patients With Suspected or Confirmed COVID-19 Study | NCT04329832 | Hydroxychloroquine<br>Azithromycin | - | Phase 2 open label, parallel design RCT | COVID Ordinal Outcomes Scale at 14 days<br>Hospital-free days at 28 days | N= 300<br>18 Yrs and older (Adult, Older Adult)<br>All sex | USA | Start- Mar 30, 2020<br>End- Dec 31, 2020 |
| 11 | Pre-exposure Prophylaxis for SARS-Coronavirus-2 | NCT04328467 | Hydroxychloroquine | Placebo | Phase 3 RCT, parallel design, quadruple masking | COVID-19-free survival<br>Incidence of confirmed SARS-CoV-2 detection | N= 3500<br>18 Yrs and older(Adult, Older Adult<br>All sex | USA | Start-Apr 2020<br>End- Aug 2020 |
| 12 | COVID MED Trial - Comparison Of Therapeutics for Hospitalized Patients Infected With SARSCoV-2 | NCT04328012 | Lopinavir/ritonavir<br>HydroxychloroquineSulfate<br>Losartan | Placebo | Phase 2<br>Phase 3 RCT, parallel design, quadruple masking | National Institute of Allergy and Infectious Diseases<br>COVID-19 Ordinal Severity Scale (NCOSS) | N= 4000<br>18 Yrs and older(Adult, Older Adult<br>All sex | USA | Start-Apr 6, 2020<br>End-Jan 1, 2021 |
| 13 | Hydroxychloroquine Versus Placebo in COVID-19 Patients at Risk for Severe Disease | NCT04325893 | Hydroxychloroquine | Placebo | Phase 3 RCT, parallel design, doublemasking | Number of death from any cause, or the need for intubation and mechanical ventilation during the 14 days following inclusion and start of treatment | N= 1300<br>18 Yrs and older(Adult, Older Adult<br>All sex | France | Start-Apr 2020<br>End- Sep 2020 |
| 14 | Proactive Prophylaxis With Azithromycin and Chloroquine in Hospitalized Patients With COVID-19 | NCT04322396 | Azithromycin<br>Hydroxychloroquine | Placebo | Phase 2 RCT, parallel design, quadruple masking | Number of days alive and discharged from hospital within 14 days | N= 226<br>Child, Adult, Older Adult<br>All sex | Denmark | Start-Apr 2020<br>End-Oct 2020 |
| 15 | Safety and Efficacy of Hydroxychloroquine Associated With Azithromycin in SARSCov-2 Virus | NCT04322123 | Hydroxychloroquine Oral Product<br>Hydroxychloroquine + azithromycin | - | Phase 3 open label RCT, parallel design | Evaluation of the clinical status Ordinal scale in 7 days | N= 630<br>18 Yrs and older(Adult, Older Adult<br>All sex | Brazil | Start- Apr 6, 2020<br>End- Aug 2020 |
| 16 | Safety and Efficacy of Hydroxychloroquine Associated With Azithromycin in SARSCoV2 Virus (Coalition Covid-19 Brasil II) | NCT04321278 | Hydroxychloroquine + azithromycin<br>Hydroxychloroquine | - | Phase 3 open label RCT, parallel design | Evaluation of the clinical status<br>All-cause mortality | N= 440<br>18 Yrs and older(Adult, Older Adult<br>All sex | Brazil | Start- Mar 28 , 2020<br>End- Aug 30, 2020 |
| 17 | Norwegian Coronavirus Disease 2019 | NCT04316377 | Hydroxychloroquine sulfate |  | Phase 4 open label RCT, parallel design | Rate of decline in SARSCoV-2 viral load | N= 202<br>18 Yrs and older(Adult, Older Adult<br>All sex | Norway | Start- Mar 25 , 2020<br>End- Apr 1, 2021 |
| 18 | Post-exposure Prophylaxis / Preemptive Therapy for SARS Coronavirus-2 | NCT04308668 | Hydroxychloroquine | Placebo | Phase 3<br>RCT,<br>parallel design,<br>quadruple masking | Incidence of COVID19 Disease among asymptomatic at trial entry | N= 3000<br>18 Yrs and older(Adult, Older Adult<br>All sex | USA | Start-Mar 17 ,<br>2020<br>End- Apr<br>21, 2021 |
| 19 | Comparison of Lopinavir/ Ritonavir or Hydroxychloroquine in Patients With Mild Coronavirus Disease (COVID-19) | NCT04307693 | Lopinavir/ritonavir<br>Hydroxychloroquine Sulfate | - | Phase 2<br>open label RCT,<br>parallel design | Viral load<br>Viral load change<br>Time to clinical improvement (TTCI) | N= 150<br>16 to 99 Yrs<br>(Adult, Older Adult<br>All sex | Republic of Korea | Start-Mar,<br>2020<br>End-May,<br>2021 |
| 20 | Randomized Clinical Trial for the Prevention of SARS-CoV-2 Infection (COVID-19) in Healthcare Personnel | NCT04334928 | Emtricitabine/tenofovir disoproxil<br>Hydroxychloroquine | Placebo | Phase 3<br>RCT,<br>parallel design,<br>Double masking | Number of confirmed symptomatic infections of SARS-CoV-2 (COVID-19) | N= 4000<br>18 Yrs and older(Adult, Older Adult<br>All sex | Spain | Start-Apr 1<br>2020<br>End-June 30<br>2021 |
| 21 | A prospective, open label, randomized, control trial for chloroquine or hydroxychloroquine in patients with mild and common novel coronavirus pulmonary (COVID-19) | ChiCTR2000030054 | Hydroxychloroquine sulfate 0.2g bid x 14 days<br>The first dose of chloroquine phosphate 1gx2 days, and the third day 0.5gx12 days | Standard of care | Prospective open label RCT | Clinical recovery time<br>Time to 2019-nCoV RT-PCR negativity in upper and lower respiratory tract specimens | N= 100<br>18 to 75 Yrs<br>All sex | China | Start-Feb 22 2020<br>End-May 5 2020 |
| 22 | A prospective, randomized, open label, controlled trial for chloroquine and hydroxychloroquine in patients with severe novel coronavirus pneumonia (COVID-19) | ChiCTR2000029992 | Chloroquine phosphate 1.0gx2 days for the first dose, 0.5gx12 day from the third day<br>Hydroxychloroquine sulfate 0.2g bid x 14 days | Standard of care | open label RCT | Clinical recovery time.<br>Changes in viral load of upper and lower respiratory tract samples compared with the baseline | N= 100<br>18 to 75 Yrs<br>All sex | China | Start-Feb 17 2020<br>End-May 5 2020 |
| 23 | Evaluation the Efficacy and Safety of Hydroxychloroquine Sulfate in Comparison with Phosphate Chloroquine in Mild and Common Patients with Novel Coronavirus Pneumonia (COVID-19): a Randomized, Open-label, Parallel, Controlled Trial | ChiCTR2000029899 | Hydroxychloroquine sulfate<br>Day1: first dose: 6 tablets (0.1g/tablet) , second dose: 6 tablets (0.1g/tablet) after 6h ; Day2-5: 2 tablets (0.1g/tablet), BID<br>Chloroquine phosphate<br>Day1-3 : 500mg, BID Day4-5: 250mg, BID | - | open label RCT, parallel design | Time to Clinical recovery<br>All-cause mortality of 28-days | N= 100<br>18 Yrs and above<br>All sex | China | Start- Feb 17 2020<br>End- Apr 30 2020 |
| 24 | Evaluation the Efficacy and Safety of Hydroxychloroquine Sulfate in Comparison with Phosphate Chloroquine in Severe Patients with Novel Coronavirus Pneumonia (COVID-19): a Randomized, Open-Label, Parallel, Controlled Trial | ChiCTR2000029898 | Hydroxychloroquine sulfate<br>Day1: first dose: 6 tablets (0.1g/tablet) , second dose: 6 tablets (0.1g/tablet) after 6h ; Day2-5: 2 tablets (0.1g/tablet), BID<br>Chloroquine phosphate<br>Day1-3 : 500mg, BID Day4-5: 250mg, BID | - | open label RCT, parallel design | Time to Clinical recovery<br>All-cause mortality of 28-days | N= 100<br>18 Yrs to 75yrs<br>All sex | China | Start- Feb 17 2020<br>End- Apr 30 2020 |
| 25 | Hydroxychloroquine treating novel coronavirus pneumonia (COVID-19): a randomized controlled, open label, multicenter trial | ChiCTR2000029868 | Hydroxychloroquine sulfate | Standard of care | open label multicenter RCT, | Viral nucleic acids<br>Adverse events | N= 360<br>18 yrs and above<br>All sex | China | Start- Feb 6 2020<br>End- June 30 2020 |
| 26 | A prospective, randomized, open-label, controlled clinical study to evaluate the preventive effect of hydroxychloroquine on close contacts after exposure to the Novel Coronavirus Pneumonia (COVID-19) | ChiCTR2000029803 | Hydroxychloroquine, small and high dose 2 arms<br>Abidol small and high dose 2 arms | - | Prospective open label RCT | Number of patients who have progressed to suspected or confirmed within 24 days of exposure to new coronavirus | N= 320<br>18 yrs to 60 yrs<br>All sex<br>Four arms total with 80 patients each | China | Start- Feb 20 2020<br>End- Feb 20, 2021 |
| 27 | Therapeutic effect of hydroxychloroquine on novel coronavirus pneumonia (COVID-19) | ChiCTR2000029559 | Hydroxychloroquine 0.1g oral 2 times/ day<br>Hydroxychloroquine 0.2g oral 2 times/ day | Placebo | Prospective placebo controlled RCT | The time when the nucleic acid of the novel coronavirus turns negative | N= 300<br>30 yrs to 65 yrs<br>All sex<br>Three arms total with 100 patients each | China | Start- Jan 31 2020<br>End- Feb 29, 2020 |

## Notes

### Competing Interest Statement

The authors have declared no competing interest.

### Funding Statement

This study was NOT funded by any institution or third party. There are no conflict of interests. No financial support was taken from any person or agency

